# Temporal Relationship Between Psychiatric Consultations and Suicide: A Postmortem Study in Osaka

**DOI:** 10.1101/2025.06.23.25330125

**Authors:** Ryu Murakami

## Abstract

This study aimed to investigate the background information of suicide cases with a history of psychiatric consultations using postmortem data from the Osaka Prefectural Medical Examiner’s Office. The goal was to explore the relationship between psychiatric consultations and the timing of suicide. Methods: A retrospective analysis was conducted on 191 suicide cases recorded in 2017. Background data such as gender, age, occupation, method of suicide, and the time interval between the last psychiatric consultation and suicide were analyzed. Descriptive statistics and correlation analysis were used to examine trends. Results: Females accounted for 55.0% of the cases, with most individuals being middle-aged (56.0%) and unemployed (70.7%). Hanging (44%) was the most common suicide method, while Mondays (24.1%) had the highest suicide frequency. The median time between the last psychiatric consultation and suicide was 9 days, and a strong negative correlation (r = -0.79) was observed between this interval and the number of suicides. Conclusion: The findings suggest that suicides often occur shortly after psychiatric consultations, emphasizing the need for timely intervention. Postmortem data can offer critical insights into suicide risks. However, further research is needed to better understand psychiatric consultation details and their link to suicide prevention.

## 1. Background

In Japan, “suicide” is treated as an unnatural death in the practice of death certification. When a death is initially determined as unnatural by the police, a detailed investigation is conducted to determine the cause. Even in cases such as deaths following transportation to medical facilities due to traffic accidents, unnatural injuries, or suicide attempts, or other external causes of death, the police conduct an examination, and once the connection between the death and a criminal act is ruled out, the body is determined to be a non-criminal death.

According to Japan’s Act on Preservation of Bodies for Postmortem Examination, when an unnatural death is deemed unlikely to have been caused by a crime based on police investigations, the medical examiner is required to perform a postmortem examination to clarify the cause of death with the aim of improving public health. In the case of suicides, Japan’s current system lacks a standardized format for recording suicide-related data across police, medical institutions, and other administrative agencies, making detailed epidemiological studies difficult. Consequently, the number of reports addressing suicides is limited.

Suicide is an urgent issue not only in Japan but also worldwide [1]. Risk factors for suicide include a history of suicide attempts, health problems, alcohol and substance abuse, and the breakdown of relationships [1]. These various factors are thought to interact, contributing to the occurrence of suicide. In particular, mental illness is known as a risk factor for suicide [2]. Epidemiological studies on suicide have been conducted in many countries, revealing findings such as a high suicide rate in Finland among patients hospitalized for depression within the first few days after discharge from psychiatric care [3]. Similarly, in England and Wales, psychiatric inpatients were most likely to die by suicide during the first week after discharge [4].

However, in Japan, there has been no research analyzing the background factors of suicide cases with a history of psychiatric consultations based on primary data collected in the field of forensic death investigation, focusing on the relationship between psychiatric consultations and the timing of suicide acts.

Therefore, this study aims to analyze the relationship between background factors related to suicide and the timing of psychiatric consultations and suicide acts, based on background information of suicide deaths.

## 2. Objective

The purpose of this study is to investigate the background information of suicide cases with a history of psychiatric consultations, based on the information recorded in the documents submitted by the police to the medical examiner when requesting a postmortem examination. Additionally, the study aims to clarify the relationship between the last psychiatric consultation date and the date of the suicide act.

## 3. Methods

### 3.1 Research Design

This study is a retrospective observational study using information from the documents sent by the police to the medical examiner when requesting a postmortem examination for suicide cases (hereafter referred to as “the target documents”). The study was conducted with the approval of the Meiji University of Integrative Medicine Human Ethics Review Committee (Approval Number: 2022-039). Approval for accessing the target documents was also obtained from the Osaka Prefectural Medical Examiner’s Office.

### 3.2 Content of the Target Documents

The target documents for this study are primary materials sent by the police to the medical examiner that record information on unnatural deaths, from which only suicide cases were extracted. The key items recorded in the target documents are as follows: ➀ Date and time of discovery, ➁ Gender, ➂ Age (recorded as an exact number), ➃ Pension status, ➄ Receipt of health and welfare services, ➅ Occupation, ➆ Co-residents, ➇ History of psychiatric consultations (including psychosomatic medicine and mental health clinics), ➈ Date of the last psychiatric consultation (recorded as a specific date), ➉ Method of suicide, ⑪ A remarks section (free text) where the police officer in charge of the investigation provides an overview of the unnatural death. Of these items, everything except age, the last psychiatric consultation date, and the remarks section is presented in a checkbox format. The authors compiled these data into a database using Excel (Microsoft Corporation, USA) and performed descriptive and bivariate statistical analyses, including correlation analysis.

### 3.3 Subjects

This study investigated 569 suicide cases recorded in the target documents that occurred in Osaka City between January 1, 2017, and December 31, 2017. In some cases of unnatural death, the body had already undergone decomposition or damage at the time of discovery, making it difficult to identify the deceased. Additionally, there were cases where the most recent living situation, occupation, or lifestyle of the deceased was still under investigation when the police sent the target documents to the medical examiner. In this study, such cases were treated as missing data, and any cases with ongoing investigations in either the evaluation items or explanatory variables were excluded from the analysis.

### 3.4 Analysis Methods

To summarize the characteristics of the subjects, continuous variables were presented as medians and interquartile ranges, while categorical variables were presented as counts and percentages. After conducting descriptive statistical analysis on the background of the suicide cases, the difference in days between the last psychiatric consultation and the date of the suicide act (hereafter referred to as “days of difference”) was calculated, and Pearson’s product-moment correlation coefficient was obtained. Based on previous studies [5], the cut-off point for the analysis of the days of difference was set at 30 days. The significance level for the tests was set at α = 0.05 (two-tailed). All statistical analyses in this study were performed using IBM SPSS Statistics Ver.29.0.0.0 (International Business Machines Corporation, USA).

## 4. Results

### 4.1 Background of Suicide Cases

Out of the 569 cases recorded during the study period, 191 cases with a history of psychiatric consultation, excluding those with missing data, were analyzed (Figure 1). The background information of the study subjects is shown (Table 1). Females accounted for 105 cases (55.0%), which was higher than the 86 cases (45.0%) of males. In terms of occupation, 135 cases (70.7%) were unemployed, the most common category. Regarding the method of suicide, hanging accounted for 84 cases (44.0%), followed by jumping from a height with 76 cases (39.8%), and poisoning with 15 cases (7.9%). The day of the week with the highest number of suicides was Monday with 46 cases (24.1%), followed by Tuesday with 30 cases (15.7%). The median number of days of difference (interquartile range) was 9 days (3-25 days). The number of suicides where the days of difference were within the 30-day cut-off was 154 cases, representing 80.6% of the total (191 cases). A scatter plot was generated using the days of difference on the X-axis and the number of suicides on the Y-axis (Figure 2).

**Table 1.** Background of Suicide Cases with a History of Psychiatric Consultations.

| Psychiatric Consultation History |  |
| --- | --- |
| n (%) | 191 (100) |
| **Sex** |  |
| Male | 86 (45.0) |
| Female | 105 (55.0) |
| **Age** |  |
| Young: under 39 years | 46 (24.1) |
| Middle-aged 40–59 years | 107 (56.0) |
| Older: 60 years and above | 38 (19.9) |
| Median (IQR<sup>a</sup>) | 53 (38–68) |
| **Occupation** |  |
| Unemployed | 135 (70.7) |
| Employed | 51 (26.7) |
| Student | 5 (2.6) |
| **Presence of cohabitants** |  |
| No | 84 (44.0) |
| Yes | 107 (56.0) |
| **Method of suicide** |  |
| Hanging | 84 (44.0) |
| Jumping from heights | 76 (39.8) |
| Jumping in front of a train | 4 (2.1) |
| Drowning | 8 (4.2) |
| Poisoning | 15 (7.9) |
| Others(e.g., use of sharp objects) | 4 (2.1) |
| **Day of the Week of Suicide** |  |
| Monday | 46 (24.1) |
| Tuesday | 30 (15.7) |
| Wednesday | 18 (9.4) |
| Thursday | 26 (13.6) |
| Friday | 27 (14.1) |
| Saturday | 24 (12.6) |
| Sunday | 20 (10.5) |

<sup>a</sup>)Interquartile Range

**Figure 1.**
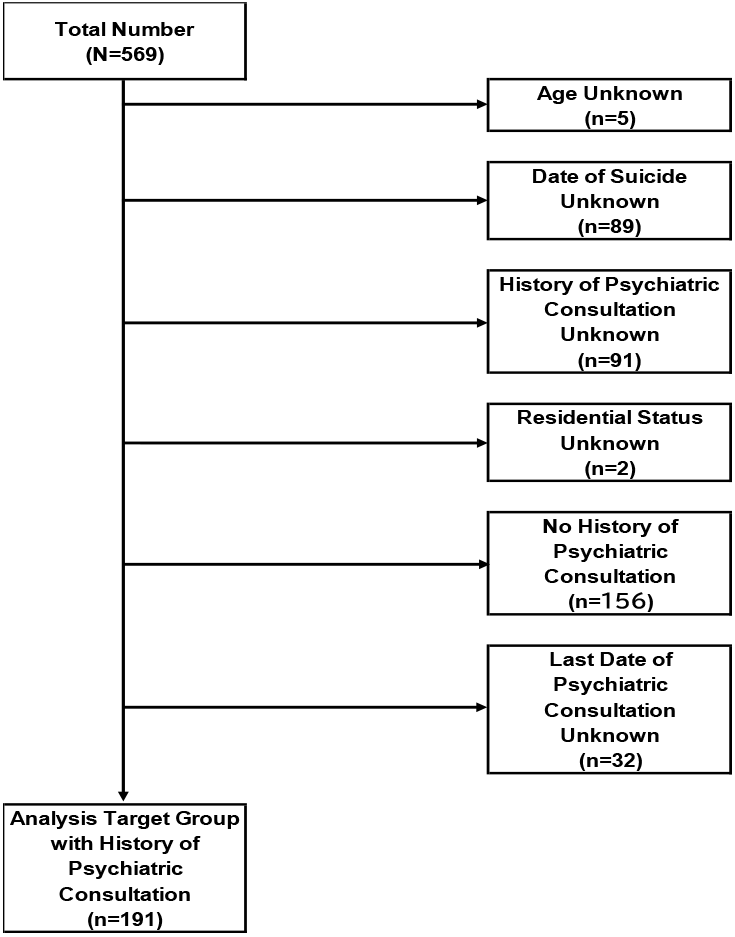
Exclusion Criteria

**Figure 2.**
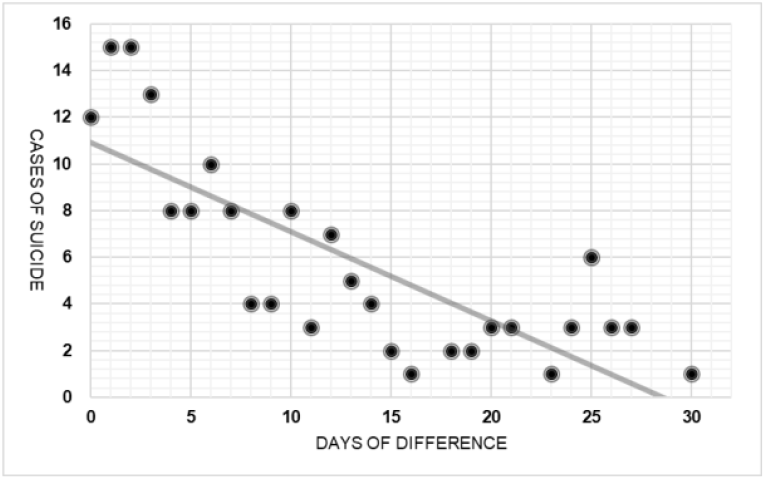
Relationship Between Days of Difference and Number of Cases

### 4.2 Relationship Between Days of Difference and Number of Suicides

For the group where suicide occurred within 30 days of the last psychiatric consultation, Pearson’s product-moment correlation coefficient was calculated to analyze the relationship between the days of difference and the number of suicides (Table 2). The correlation coefficient was -0.79 (95% CI; -0.90 to -0.58), indicating a strong negative correlation.

**Table 2.** Relationship Between Days of Difference and Cases of Suicides.

|  | Correlation Coefficient | 95%CI | p |
| --- | --- | --- | --- |
| Difference in Days / Number of Cases | -0.79 (-.90~-0.58) |  | <0.01 |

## 5. Discussion

### 5.1 Background of Suicide Cases

In the breakdown of suicide cases (Table 1), the proportion of females was higher. According to previous studies analyzing factors associated with a history of psychiatric consultations among completed suicide cases, being female was identified as a factor related to having a history of psychiatric consultations [6]. Kodaka et al. also found that the rate of self-harm and previous suicide attempts was significantly higher among females compared to males [7]. The higher proportion of females in this study suggests a possible relationship between the higher rate of self-harm in females and having a history of psychiatric consultations, as seen in previous studies. Further investigation into the relationship between self-harm and access to mental health care is needed.

In terms of age distribution, younger and older age groups were less represented compared to the middle-aged group. Previous studies on individuals who died by suicide without receiving mental health services reported that younger and older individuals were more likely to not use mental health services. Research on barriers and facilitators for seeking mental health support among young people [8] indicated that low mental health literacy and the stigma associated with seeking help from others were barriers to accessing mental health care. Similarly, in this study, the lower representation of older adults may suggest that difficulties in accessing mental health care influenced the results. For older adults, compared to younger individuals, it is likely more difficult to access mental health information through the internet or smartphones. A scoping review on the barriers and facilitators to the use of e-health services by older adults found that low self-efficacy, lack of knowledge, and insufficient support regarding the benefits of e-health services were barriers to their use [9]. Additionally, a lack of awareness of the need for mental health care and fear of hospitalization have been identified as major barriers to accessing care [10]. Therefore, active outreach strategies promoting mental health care for both younger and older individuals may be essential for suicide prevention.

The breakdown by occupation revealed that the unemployed group had the highest proportion. It is assumed that unemployed individuals have fewer opportunities for social interaction through work or school compared to employed individuals or students. Previous studies have suggested that social isolation may be a risk factor for suicide [11]. The findings of this study involve cases where individuals ultimately died by suicide despite receiving mental health care prior to their deaths. Based on these results, it is important to evaluate the intensity of mental health care interventions for high-risk groups and to implement continuous and proactive preventive interventions involving family and governmental support following psychiatric consultations.

Regarding the day of the week, suicides occurred most frequently on Mondays compared to other days. Since Monday is typically the first day after a weekend, it is possible that psychological stress related to returning to work or school was a contributing factor. Previous studies have suggested that Mondays and seasons may be factors related to an increase in the number of suicides [12, 13]. On the other hand, a study examining suicides matched by age and gender found no evidence that deaths by suicide were more frequent on Mondays compared to other days [14]. Taken together, these reports indicate that further investigation is needed into the relationship between days of the week and suicidal behavior. Several studies have explored the relationship between life cycles and the frequency of suicides, suggesting a potential causal relationship between the two. However, since the unemployed group was the largest in this study and the relationship between the day of the week and occupation was not examined, it is necessary to consider the possibility of unknown factors contributing to the increase in suicides on Mondays.

In terms of suicide methods, hanging and jumping were more frequent than other methods. Since this study focused on cases where individuals died by suicide, it is possible that methods with higher lethality were more common. Reports investigating the lethality of suicide methods indicate that firearms have the highest lethality rate, followed by hanging and drowning [15]. In Japan, strict regulations on the possession and use of firearms significantly limit the general public’s access to them, with only certain law enforcement personnel permitted to possess and use firearms under specific circumstances. Therefore, it is likely that deaths by suicide in Japan are carried out using more accessible methods other than firearms. Suicide rates by method vary across countries [16], with regional differences in the regulation of items such as firearms and drugs potentially playing a role. It is essential to both regulate high-risk locations and methods as much as possible and to identify high-risk individuals early, implementing active preventive interventions.

### 5.2 Relationship Between Days of Difference and Number of Suicides

A strong negative correlation was found between the days of difference and the number of suicides, as indicated by the Pearson’s product-moment correlation coefficient. Several studies investigating suicides among patients discharged from psychiatric hospitals have shown that the highest suicide rates occur immediately after discharge [3] [5] [4]. In this study, data on the presence or absence of psychiatric consultations were entered based on checkboxes in the primary documents, so the specific type of consultation, such as inpatient or outpatient, was not recorded. However, similar to findings in international studies, there appears to be a relationship between psychiatric consultations and the risk of suicide in Japan as well.

Since psychiatric consultations generally occur when individuals recognize symptoms, either subjectively or through others’ observations, it can be inferred that the suicides in this study may have occurred during times when psychiatric symptoms were most pronounced, leading to the decision to seek psychiatric care. Future research should investigate the background of psychiatric consultations, including details such as inpatient vs. outpatient status, first-time vs. follow-up consultations, and initial diagnoses at the first consultation. Regardless, this study provides a new perspective, as it is based on primary data obtained from a forensic viewpoint in Japan, and continued investigation will be necessary.

## 6 Conclusion

Using postmortem data collected from a forensic perspective, this study examined the background information of suicide cases with a history of psychiatric consultations. The background characteristics of those who died by suicide included a higher proportion of females, middle-aged individuals, the unemployed, and those living with others. Regarding the circumstances of the suicides, a higher proportion occurred on Mondays and by hanging. The number of days between the last psychiatric consultation and the suicide act indicated that suicides tended to occur within a short period after the psychiatric consultation.

In Japan, the analysis of suicide cases using postmortem data is still in the stage where active data collection is necessary. Continued investigation of postmortem data has the potential to provide a foundation for suicide prevention efforts.

## Limitations

This study’s data were constructed based on the information available at the time when the police had inquired with medical institutions and when the target documents were received by the medical examiner. Therefore, it is possible that some information was unknown at the time the documents were submitted to the medical examiner. Additionally, there may be confounding factors that were not included in the dataset used in this study, which must be taken into consideration.

The following research limitations should be noted: First, this study only focused on individuals who completed suicide, and thus, does not examine cases of attempted suicide. Second, cases involving multiple methods of suicide, such as taking medication followed by hanging, could not be tracked.

## Data Availability

The data used in this study were obtained from official documents archived at the Osaka Prefectural Medical Examiner's Office. Due to the sensitive nature of the information and institutional restrictions, these data are not publicly available and cannot be shared. All materials were anonymized prior to analysis to ensure that no individuals could be identified.

## Data Availability Statement

The information on suicides used in this study is based on documents stored at the Osaka Prefectural Medical Examiner’s Office. Therefore, this information is not publicly available. The materials used in this study were processed to ensure that individuals could not be identified. All statistical analyses were performed on a secure, offline information terminal, and the data used were stored on encrypted storage media, which is strictly managed within the lead author’s affiliated research institution.

## Ethics Statement

This study was conducted following an opt-out procedure. The opt-out procedure was created in accordance with ethical guidelines in Japan, allowing the families of the deceased to freely request exclusion from this study. The opt-out document is published on the website of the lead author’s affiliated research institution. A printed version of this document is also posted in a designated location within the author’s institution and is available upon request.

## Acknowledgments

We would like to express our gratitude to the Osaka Prefectural Medical Examiner’s Office for providing the data used in this study. This study was supported by JSPS KAKENHI (Grant Number: JP22K21128), Grant of The T. and F. Kitamura Foundation for Studies and Skill Advancement in Mental Health (Grant Number: K2023-0004), and a research grant from Meiji University of Integrative Medicine (Grant Number: MUIM-B-9).

